# Fluency Changes due to Sports-Related Concussion

**DOI:** 10.1101/2021.09.19.21263791

**Authors:** Sona Patel, Caryn Grabowski, Vikram Dayalu, Mercedes Cunningham, Anthony J. Testa

## Abstract

**Background:** Alterations in speech have long been identified as indicators of various neurologic conditions including traumatic brain injury (TBI), neurodegenerative diseases, and stroke. TBIs that can be assessed using the Glasgow Coma Scale often result in speech symptoms such as dysarthria and occasionally neurogenic stuttering. The manifestation of symptoms including the specific changes in speech occurring in mild TBIs (or concussions) may differ from more severe head injuries. This work aims to compare speech fluency in sport-related concussion to baseline performance as well as non-athlete controls.

**Methods:** A total of 230 Division I student athletes participated in pre-season speech testing. Of these, 12 students (18-22 years) who sustained a concussion also participated in speech testing in the days following diagnosis of concussion. Samples of picture descriptions were independently coded by three trained raters as 17 error types within the three traditional categories of errors defined in fluency analysis (Stuttering-Like Disfluency, Articulation Error, Other Disfluency).

**Results:** Within-subjects analysis comparing the difference in percent error scores at baseline and post-concussion revealed significant differences for interjections (t(11)=-2.678, p< .05). The Other Disfluency category was also significantly different (t(11)= -2.735, p< .05), with more errors occurring after a concussion. No change in the Stuttering-Like Disfluency (t(11)= -0.799, p>.05) or Articulation Error category (t(11)=-0.045, p>.05) was found.

**Conclusions:** These results demonstrate that speech changes occur following mild sports-related concussions. Specifically, the rate of interjections increased in a limited sample of college athletes who sustain a concussion. Changes in additional error types (fillers, pauses) were trending, but were not significant potentially due to the low sample size. Future studies should consider speech as a diagnostic tool for concussion.

## I. INTRODUCTION

Alterations in speech production are widely accepted as hallmark sequelae of neurotrauma resulting from traumatic brain injury, stroke, and other neurologic conditions (Yorkston, 1996). A traumatic brain injury or TBI is defined as an insult to the brain in a “closed head” nature due to forces of acceleration and deceleration on impact with subsequent consequence to neurochemical and neurometabolic functions (Signoretti et al., 2011). Although there is some variability in the usage of the terms “TBI” and “concussion”, the term concussion is often used to refer to the mild forms of TBI (Mayer et al., 2017). While often not visible with neuroimaging, a concussion can result in a host of symptoms that may include physical, psychosocial, and cognitive components (Ryan & Warden, 2003).

Often, detection of concussion relies on a series of assessments focusing on measures of physical symptoms (e.g., balance, visual disturbances, headache, fatigue) as well as some basic cognitive functions (e.g., concentration, recall) (Echemendia et al., 2017; Broglio et al., 2014). Such symptoms are paralleled to more dramatic presentations in TBIs where significant and persistent impairments in physical and cognitive functions are widely recognized. In addition to these symptoms, more severe head injuries are also known to result in speech disturbances impacting total communication. The most common of these is dysarthria (Wang et al., 2005; Kuruvilla et al., 2012), or impairment in strength, tone, and/or coordination which may result in trace to severe alterations in articulatory precision, vocal quality, rate, and prosody. Other speech problems include apraxia of speech (Duffy, 2016) and occasionally neurogenic stuttering (Norman et al., 2018), resulting in disruptions in motor sequencing, smoothness, rate, and other prosodic qualities in speech production.

Despite the relative parallels in physical and cognitive symptoms identified in concussion and more severe head injuries, exploration of the impacts of *concussion* on speech production has been limited. Changes in speech are typically not captured on commonly used symptom inventories for milder injuries and sports-related sideline assessments (Schatz et al., 2006; Asken et al., 2020). However, early evidence has shown significant alterations in rate of speech (Salvatore et al., 2019) and articulatory precision (Chong et al., 2021) in the presence of concussion. Recent case reports of disturbances in fluency have also been reported (Robertson & Diaz, 2020; Rose et al., 2021; Toldi & Jones, 2021). Although such reports document more pronounced disruptions in fluency, alterations in cognition are known to occur in concussion, affecting attention, memory, and executive functions (Karr et al., 2014). These cognitive impairments may result in reduced availability of resources to support the linguistic processing required for speech production (Corley & Stewart, 2008). These preliminary findings suggest that further examination of speech changes in milder head injuries is warranted at this time. Because error rates (disfluencies, misarticulations, speech errors) in typical speech production are low (around 6%; Fox Tree, 1995), small deviations from normal that might occur after a concussion may not be noticeable or identified as disordered because the errors do not interfere with functional communication even though these errors may be systematically or consistently occurring.

In the present study, we analyzed speech samples obtained in the days immediately following a concussion (Shriberg, 2001) and compared these samples to baseline recordings obtained prior to head injury. A picture description task was used to obtain a more ecological valid assessment of speech errors and disfluencies present. Traditional methods for examining speech errors use the fluency classification system for coding errors that occur in connected speech output. These include three general categories, stuttering-like disfluencies (e.g., prolongations, repetitions), articulation errors (e.g., insertions, deletions) and other disfluencies (e.g., pauses, fillers) (Ambrose & Yairi, 1999). Analysis involves tallying the frequency totals by type of error and in total. This results in a quantified severity of speech errors/dysfluencies but also a descriptive profile of the nature of error types.

## II. METHODS

### A. Subjects

A total of 230 student athletes (18-22 years) at Seton Hall University participated in pre-season speech testing as part of this study in addition to routine baseline testing. Of the individuals tested, 12 students (4 males, 8 females; mean age: 19.9 years) experienced a concussion as determined by the team’s Certified Athletics Trainer. All subjects were proficient in English and reported having normal hearing and vision at baseline in addition to having no history of a hearing, speech, language, neurological, mood, or attention disorder. All injured subjects performed follow-up speech testing within 48 hours after symptoms were reported (the primary symptom in 10 subjects included headache/head pressure, irritability in 1 subject, and noise sensitivity in 1 subject) in aims of capturing presence of symptoms whenever they emerge, which at maximum resulted in test times of within 96 hours after the head injury occurred. All subjects signed informed consent approved by the Hackensack Meridian Health Institutional Review Board on behalf of Seton Hall University.

### B. Procedures

Participants performed a series of speech tasks during baseline and post-concussion testing in a within-subjects design. Here we report on speech obtained using a picture description task (Shimada et al., 1998). Participants were instructed to “Take a look at this picture and tell me what you see. I know there is a lot happening in this picture, so take your time before you start. When you’re ready, explain to me what’s going on in this picture.” Recordings were made using an AKG head-worn microphone (HARMON International, Stamford, CT) that was routed through an Apollo audio interface (Universal Audio, Inc., Scotts Valley, CA) before being recorded onto the computer with Adobe Audition software.

### C. Analyses

All sound files were reviewed for any technical issues and segmented to exclude any initial “Okay” that signaled acknowledgement of the task instructions and “That’s about it” or similar phrase to indicate that the subject was done speaking. All 24 sound files (12 baseline and 12 post-concussion) were transcribed for fluency analysis. Fluency analysis was independently conducted by three of the co-authors (VD, CG, SP) who are trained in performing fluency analysis. Each coding discrepancy was reviewed as a group and consensus was obtained. The raters were blinded to subject and condition when performing their evaluations. The raters identified and coded errors into 1 of 17 error types. These error types were also summed within three categories: stuttering-like dysfluencies (part-word repetition, single-syllable whole-word repetition, prolongation, block), articulation errors (substitution, distortion, addition, omission), and other dysfluencies (multisyllable whole-word repetition, revision, abandoned or incomplete segment, interjection, filler, phrase repetition, pauses) (Ambrose & Yairi, 1999; Roberts et al., 2009; Lutz & Mallard, 1986). For each sample, error totals were divided by the number of syllables produced in the sample in order to normalize the values to the amount of speech produced. This resulted in percent error scores for each error type. Since this study involved a within-subjects design to examine changes in multiple independent variables, paired samples t-tests were computed in SPSS to compare the difference in baseline percent error scores and post-concussion percent error scores for each of the 17 categories.

## III. RESULTS

On average, the number of syllables produced increased after a concussion (mean or M=89.2; standard deviation or SD=35.5) compared to baseline (M=99.1; SD=42.9), however this difference was not significant (t(11)= -0.846, p > .05). Results of paired samples t-tests of the percent error scores for all error types at baseline compared to post-injury are shown in Table 1 below. Significant differences were found for one error type, namely interjections (t(11) = -2.678, p < .05).

**Table 1.**
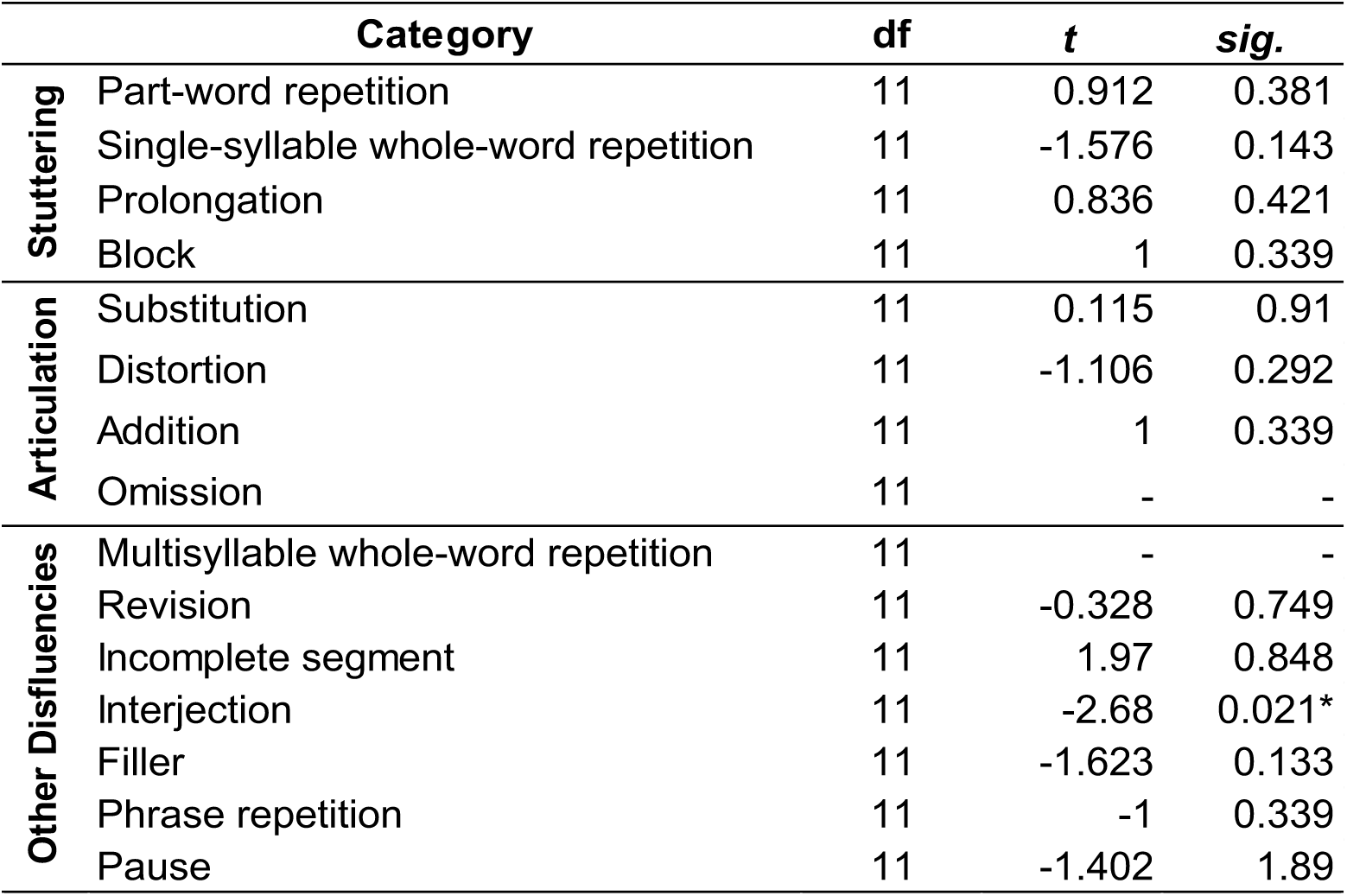
Results of paired samples t-tests of percent error scores at the α = .05 level for Baseline compared to Concussion. There were no occurrences of multisyllabic whole word repetitions and omissions for any subject.

|  | Category | df | *t* | *sig.* |
| --- | --- | --- | --- | --- |
| **Stuttering** | Part-word repetition | 11 | 0.912 | 0.381 |
|  | Single-syllable whole-word repetition | 11 | -1.576 | 0.143 |
|  | Prolongation | 11 | 0.836 | 0.421 |
|  | Block | 11 | 1 | 0.339 |
| **Articulation** | Substitution | 11 | 0.115 | 0.91 |
|  | Distortion | 11 | -1.106 | 0.292 |
|  | Addition | 11 | 1 | 0.339 |
|  | Omission | 11 | - | - |
| **Other Disfluencies** | Multisyllable whole-word repetition | 11 | - | - |
|  | Revision | 11 | -0.328 | 0.749 |
|  | Incomplete segment | 11 | 1.97 | 0.848 |
|  | Interjection | 11 | -2.68 | 0.021* |
|  | Filler | 11 | -1.623 | 0.133 |
|  | Phrase repetition | 11 | -1 | 0.339 |
|  | Pause | 11 | -1.402 | 1.89 |

An examination of the distribution of errors in the three major categories (stuttering-like dysfluencies or “SLDs”, articulation errors or “Artic”, other disfluencies or “ODs”) revealed that most errors occur within the OD category both prior to and after injury (see Figure 1). Paired samples t-tests of the SLDs, Artic errors, and ODs showed significant differences for the ODs category at the α= .05 level (t(11)= -2.735, p = .019) with more ODs occurring after a concussion. No change in SLDs (t(11)= -0.799, p = .441) or Artic errors (t(11)= -0.045, p = .965) was found. Percent error scores for each participant are shown in Figure 2 for the OD category.

**Figure 1.**
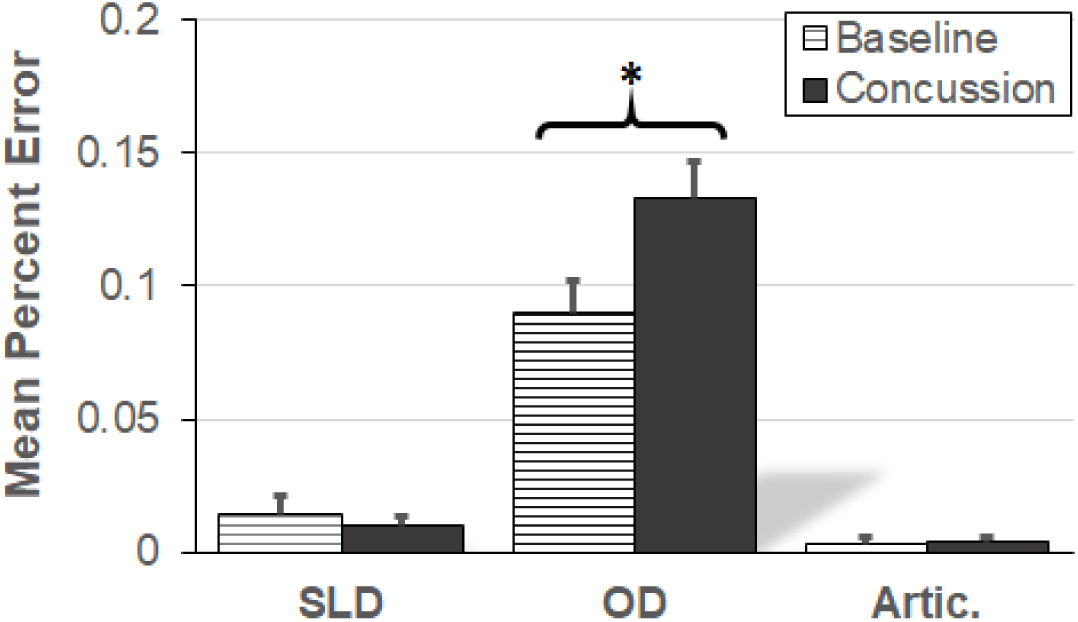
Mean percent error for stuttering-like dysfluencies (SLDs), articulation errors (Artic.), other disfluencies (OD) at baseline and after concussion. Significant paired samples t-tests reported at the α = .05 level.

**Figure 2.**
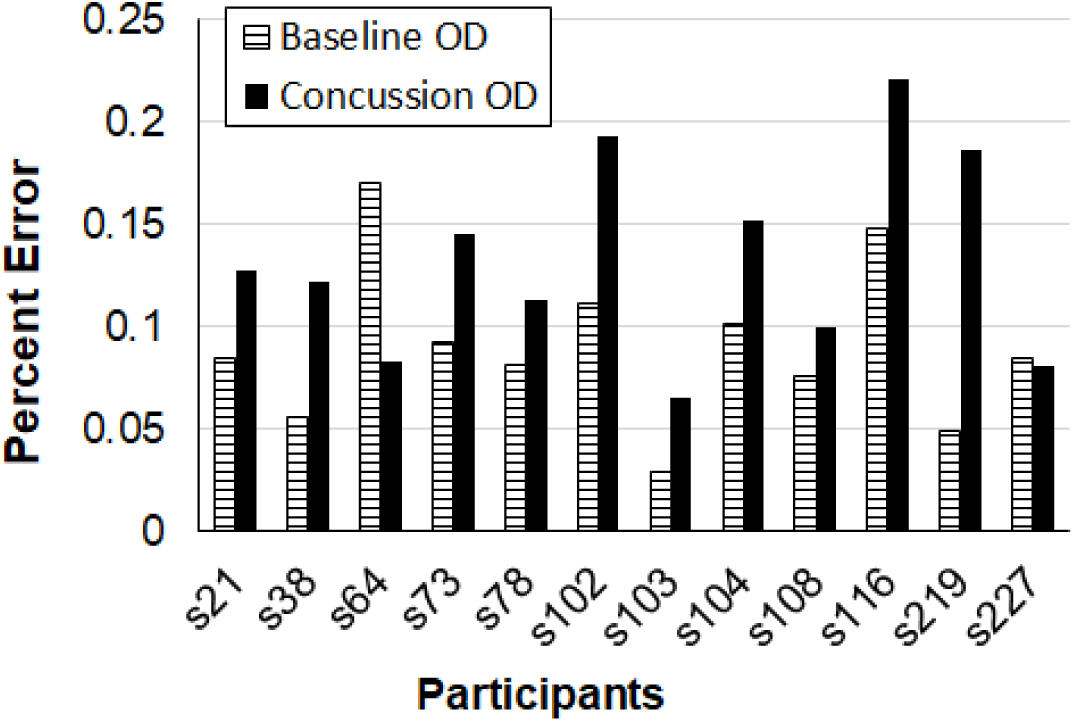
Percent error for the other disfluencies (OD) category at baseline and after concussion for each participant.

## IV. DISCUSSION

This study examined speech error patterns in Division I college athletes in the acute time period following a sports-related concussion with a prediction that a head injury may affect cognitive-linguistic functions contributing to fluent speech output, thereby impacting the extent of speech errors that occur (López-de-Ipiña et al., 2020). Baseline measures of speech disfluency in connected speech were compared to performance within 48 hours after symptoms appeared (up to 4 days following a concussion). A strength of this work is the available baseline comparison data. In notion of “normal” speech exhibiting some degree of speech errors, the analysis of pre-injury compared to concussion allows identification of discrete error trends possibly not noted as symptomatic in functional speech. As predicted, pre-post comparisons demonstrated an overall increase in total number of speech errors after a concussion (Mean Baseline = 11.5 errors; Mean Concussion = 15.9 errors), however this difference was not significant (t(11)= -1.772, p = 0.104) potentially due to the low sample size.

Of the 17 types of speech errors examined, the occurrence of ODs increased significantly after a concussion, as opposed to SLDs or articulation errors. The interjection error type was the greatest contributor to the OD error rate, increasing post-injury in all but one participant. Interjections are defined as additional words or phrases that do not contribute to the sentence structure or meaning (e.g., so, you know, like) (Ambrose & Yairi, 1999). This pattern of increased errors within the OD category has been observed in other acquired neurological conditions, such as severe TBI, adults with developmental cluttering, and Parkinson’s disease (Power et al., 2020; Myers et al., 2012; Smith et al., 2018). Both Power and colleagues (2020) and Smith and colleagues (2018) have demonstrated in different populations that such errors are associated with cognitive deficits. Concussion is known to impact cognition, specifically executive functions, attention, and memory (Covassin & Elbin, 2012) that may result in the increased rate of interjections and other similar dysfluency types (e.g., fillers, pauses).

Interestingly, the differences after concussion for the OD category were more significant than the interjection error type alone. A closer look at the changes in the errors within the OD category revealed several error types that approached significance, including fillers (e.g., “uh” and “um”) and pauses (e.g., duration longer than 250 ms). Fillers differ from interjections in that they are extraneous sounds or non-words (Corley & Stewart, 2008), while interjections are extraneous words or phrases. Although they both functionally serve to maintain continuity of connected speech production while accommodating increased demands on planning intended speech (Clark & Tree, 2002), the production of non-words has traditionally been thought of as a lower level of language processing compared to “real” words (Boles, 1997; Bose & Buchanan, 2007). However, some studies argue that fillers such as “um” have been included in regular vocabulary and thus can be considered to be categorically included with interjections (Corley & Stewart, 2008; O’Connell & Kowal, 2005). It is possible that differences in these categories are more apparent in people who are more severely injured, but for mild cases, it might be worth combining these two categories as a single variable for assessment of speech for mild concussion.

Our results showed only a trending difference in the number of pauses (minimum pause length 250 ms) after concussion, despite early evidence reporting such a difference (Banks et al., 2020). It is important to note that while reviewing speech samples, clinical presence of slowed rate of speech and increased pausing or latency between and within utterances was subjectively observed. Future work should re-analyze this data with a larger sample size. In addition, an examination based on different subtypes of pauses may reveal the presence of changes in pausing after a concussion.

Acquired neurogenic stuttering resulting from injury to the brain can arise due to damage in the brainstem, subcortical, and cortical regions of the brain (Jokel et al., 2007). Several studies suggest that, although rare, an increased rate of acquired stuttering occurs in individuals with TBI. Some studies report stuttering-like behaviors consistent with psychogenic stuttering, including pauses, speech hesitations, brief blocks, rapid repetitions, and occasional prolongations (Roth, 2015), while others find less typical disfluencies such as repetitions as the primary error type in cases of neurogenic stuttering after TBI in addition to the more common disfluencies such as interjections, silent pauses, broken words, revisions and starters (Lundie et al., 2014).

One of the limitations of this study is the inclusion of the mildest of mild cases. In order to provide the best care for student athletes, the health team identifies all possible cases of concussion that might have occurred. In other words, anyone who has taken a blow to the head undergoes sideline testing for symptoms of concussion. Speech evaluations were performed on all such cases, which may have resulted in a few referrals where symptoms of concussion were minimal. In the future, these will be controlled for by setting a minimum symptom score as part of the inclusion criteria.

## V. CONCLUSIONS

The purpose of this study was to determine whether changes in speech error patterns exist in the acute time period following a sports-related concussion compared to pre-season baseline measures. Our findings suggest the presence of increased speech errors with concussion in a limited sample of college athletes from one institution. Specifically, increased rates of interjections were observed post-head injury. Therefore, disfluencies that are not typically considered features of stuttering appear to be important when characterizing the overall speech patterns of patients with TBI. The OD category was in fact more significant than the interjections error type alone, despite the finding that other error types within the OD category were not significant. This suggests that there may be a tradeoff in how errors manifest (as interjections, pauses, revisions, fillers, etc.) and/or are assessed. We anticipate that a larger sample size would strengthen statistical findings and more appropriately determine the nature of speech errors in concussion. Taken together, these results support the need for further examination of other metrics of speech production which may serve to indicate the presence or characterize the nature of concussion or milder forms of traumatic brain injury.

## Data Availability

In compliance with IRB guidelines, deidentified data in the form of an excel sheet will be made available upon request after the project has ended.

## Ethics Approval

This study was approved by the Institutional Review Board at Hackensack Meridian Health in partnership with Seton Hall University.

## Acknowledgements

The authors would like to thank the athletics trainers Catherine Lass, Kaitlin Kelly, and Nicholas Schulman for their help and support in coordinating student athlete testing, and the student athletes for their time and efforts in participating in this study.

